# Social contact patterns following the COVID-19 pandemic: a snapshot of post-pandemic behaviour from the CoMix study

**DOI:** 10.1101/2023.08.29.23294767

**Authors:** Christopher I Jarvis, Pietro Coletti, Jantien A Backer, James D Munday, Christel Faes, Philippe Beutels, Christian L. Althaus, Nicola Low, Jacco Wallinga, Niel Hens, W John Edmunds

## Abstract

**Background:** The COVID-19 pandemic led to unprecedented changes in behaviour. To estimate if these persisted a final new round of the CoMix survey was conducted in four countries at a time when all societal restrictions had been lifted for several months.

**Methods:** We conducted a survey on a nationally representative sample in the UK, Netherlands (NL), Belgium (BE), and Switzerland (CH). Participants were asked about their contacts and behaviours on the previous day. We calculated contact matrices and compared the contact levels to a pre-pandemic baseline to estimate R_0_.

**Results:** Data collection occurred from 17 November to 7 December 2022. 7,477 participants were recruited. Some were asked to undertake the survey on behalf of their children. Only 14.4% of all participants reported wearing a facemask on the previous day, varying between 6.7% in NL to 17.8% in CH. Self-reported vaccination rates in adults were similar for each country at around 86%. Trimmed mean recorded contacts were highest in NL with 9.9 (95% confidence interval [CI] 9.0 to 10.8) contacts per person per day and lowest in CH at 6.0 (95% CI 5.4 to 6.6). The number of contacts at home were similar between the countries. Contacts at work were lowest in the UK (1.4 contacts per person per day) and highest in NL at 2.8 contacts per person per day. Other contacts were also lower in the UK at 1.6 per person per day (95% CI 1.4 to 1.9) and highest in NL at 3.4 recorded per person per day (95% CI 4.0 to 4.0). Using the next-generation approach suggests that R_0_ for a close-contact disease would be roughly half pre-pandemic levels in the UK, 80% in NL and intermediate in the other two countries.

**Conclusions:** The pandemic appears to have resulted in lasting changes in contact patterns that would be expected to have an impact on the epidemiology of many different pathogens. Further post-pandemic surveys are necessary to confirm this finding.

## Background

Pandemics do not end with a bang [1, 2] but if you’ve seen one pandemic, then you’ve seen one pandemic [3]! The much-desired return to normality following the COVID-19 pandemic was always going to be difficult to determine both in what it means and when, if ever, it might happen. The expectation that things will be the same as before is also complicated by the pandemic leaving an indelible mark on society. The demonstration for the capacity of remote working, where possible, may mean the number of people in offices will be lower. Socialising when ill could become taboo. Facemasks may become routine for some. Sentiments towards vaccines, perhaps more complex. One way we can assess the return is by conducting contacts surveys [4] to measure who mixes with whom.

During the pandemic, the CoMix study recorded epidemiologically relevant (i.e. face-to-face) social interactions in representative samples of individuals from a number of European countries (21 countries in total collected data as part of the project) [5–11]. Different countries collected data at different points during the pandemic[12–15]. However, the UK, Netherlands and Belgium initiated their surveys during the first lockdowns in Spring 2020 and collected data more or less continually for about two years. Switzerland collected data between January 2021 and May 2022. The surveys were used to provide rapid insights on how social contact behaviour adapted as a result of the pandemic and the restrictions that governments put in place. Data collection was wound up at different times, the Netherlands stopped in September 2021 and the rest of the countries in this study all stopped their CoMix surveys by Spring or early Summer 2022, as pandemic-specific restrictions were being lifted across Europe.

In this study, we return to measure epidemiologically relevant social contacts during late November and early December 2022 in the UK, Netherlands, Belgium, and Switzerland, using identical methods as for the main CoMix study. Moreover, we provide quantitative estimates of contact patterns some months after all restrictions were lifted. We compare estimates of contact patterns in this post-pandemic period (in which high rates of infection with Omicron subvariants as well as other respiratory infections was relatively common) with those measured prior to and during the pandemic. We compare the levels of mixing across the four countries and in different settings. We may not yet be at a stable post-pandemic period of behaviour, with adaptations still to come, but this study provides a bridge between how we behaved during 2020, the acute phase of the COVID-19 pandemic, and the evolving picture of where we might be heading in the years to come.

## Methods

### Ethics Statement

Participation in this opt-in study was voluntary, and all analyses were carried out on anonymised data. The study was approved in the UK by the ethics committee of the London School of Hygiene & Tropical Medicine Reference number 21795. The study to collect CoMix data in Belgium was approved by the Ethics Committee of UZA with reference 3236 - BUN B3002020000054. The Medical Research Ethics Committee (MREC) NedMec confirmed that the Medical Research Involving Human Subjects Act (WMO) does not apply to the CoMix study in the Netherlands (research protocol number 22/917). Therefore an official approval of this study by the MREC NedMec is not required under the WMO. The study to collect CoMix data in Switzerland was approved by the ethics committee of the Canton of Bern (project number 2020-02926).

### Study design

We conducted an online behavioural survey called CoMix where individuals recorded details of direct contacts in the day prior to the survey. We defined a direct contact as anyone who was met in person and with whom at least one word was exchanged, or anyone with whom the participants had any sort of skin-to-skin contact. Contacts of individuals under the age of 18 were collected by asking parents to answer on behalf of their child.

The design of the CoMix survey is based on the POLYMOD contact survey. The POLYMOD survey was a self-administered paper survey in the form of a daily diary recording participants’ social contacts [16]. In the CoMix study, participants consented to self-report their social contacts made on the day prior to survey participation. Other survey questions in CoMix included participants’ work attendance, self-reported risk status, use of facemasks, presence of recent symptoms, and vaccination history. Details of the CoMix study including the protocol, methodology, and survey instrument have been published previously [5, 8, 17].

CoMix was conducted in 21 European countries between March 2020 and July 2022. In this paper, we present the final additional round of data collected between Nov 2022 and Dec 2022 in the UK, the Netherlands, Belgium, and Switzerland. In each study country, a nationally representative sample was recruited using quota sampling based on age, gender, geographic region, and where possible, socioeconomic status to reflect the distribution within the national population. The market research company Ipsos recruited participants through a combination of social media, web advertising, and email campaigns to meet quotas.

### Study participants

The final round of CoMix ran from 17 November 2022 to 7 December 2022. Data was collected at similar times for all countries; starting first in the UK (17 Nov to 29 Nov), then the Netherlands (21 Nov to 3 Dec), Switzerland (22 Nov to 7 Dec), and finally Belgium (23 Nov to 5 Dec). As per prior rounds of CoMix and due to differing funding levels, the UK panel was double the size of the other countries with 2,991 participants (Netherlands 1,491, Switzerland 1,495, Belgium 1,500). Most of the data was collected in adults, with a proportion of parents reporting on behalf of their children.

### Data

#### Reporting of contacts

The participants reported their contacts from the day prior to the survey in two ways: individual contacts and group contacts. Individual contacts were recorded by asking the participant to list each contact and their characteristics separately. Following this, we asked whether they had recorded all their contacts. If they had not, then they provided details of the total number of contacts they had at work, school, or other settings for the age groups 0 to 17, 18 to 59, and 60+, both overall and for physical contacts only (‘group contacts’). They were also asked how often they met each contacted person, how much time was spent with them, and their relationship with the contacted person. Further details of the CoMix survey have been reported extensively previously [5, 6, 8, 9].

#### Demographic information

The survey captures information about participants’ demographics. Participants’ ages were grouped into categories of 0-4, 5-11, 12-17, 18-29, 30-39, 40-49, 50-59, 60-69, and 70 years and above. Participants were asked to report how they describe their gender, with the options of “Female”, “Male”, “In another way”, or “Prefer not to answer”. Participants were also asked about their household size.

#### Risk perception, status, and mitigation

Participants reported about their uptake of risk mitigating activities and responded to statements regarding their perception of risk. Participants were asked to rate the following statements: (i) “I am likely to catch coronavirus”; (ii) “I am worried that I might spread coronavirus to someone who is vulnerable”; and (iii) “Coronavirus would be a serious illness for me” with the Likert scale of “Strongly agree,” “Tend to agree”, “Neutral”, “Tend to disagree” and “Strongly disagree”. Participants self-reported whether they considered themselves to be high risk, whether they wore a face covering at least once on the prior day, and their COVID-19 vaccination status.

#### Presentation of COVID-like symptoms

Participants reported COVID-19-compatible symptoms in the 7 days prior to survey participation. These symptoms included: fever or chills, cough, shortness of breath (or difficulty breathing), fatigue (or extreme tiredness), muscle or body aches or headache, congestion (or runny nose), and sore throat.

#### Work status and attendance

Participants were asked to report if they were employed, and if so, whether they were full time, part time, or self-employed. They reported whether their work place was open and whether they attended work on the day prior to responding to the survey (the day on which they reported contacts for).

### Statistical analysis

R version 4.1.1 [18] was used for all analyses, and the code and data are available online (see Data Availability Statement). The code for the analyses conducted in this study are available on https://github.com/jarvisc1/cmix_post_pandemic.

### Descriptive

We calculated the counts and percentages for contacts, risk perceptions, mitigations, symptoms, and employment related questions stratified by age, gender, household size, day of the week. While parents answer as proxies for children in the study, we describe the designated child as the “participant” where applicable. We restricted the analysis to adults only for risk perception, mitigation, symptoms, and employment questions, as we consider the data to be more reliable than those reported for children by their parents. For risk perception, we present the number and percentage of adults who strongly agreed with the statements asked.

### Mean number of contacts

We calculated the mean number of contacts for each of the characteristics presented in the descriptive analysis. We used a cut-off value of 100 as the maximum for contacts. This means we counted any individual who reported more than 100 contacts as if they reported 100 contacts to reduce the weight of individuals reporting high numbers of contacts on the mean. Previous publications, specifically for the UK papers for CoMix have used a cut-off of 50 [8]. The value of 100 was chosen for two reasons, 1) Over 99.9% of participants reported contacts of less than 100, 2) The previous publication of CoMix comparing 21 countries [17] used a cutt-off of 100, so for sake of consistency we used this threshold. For mean contacts by setting and country we calculated 95% confidence intervals (95% CI) using bootstrapping, similar to the approach used in a previous CoMix publication [17]. For mean contacts by characteristics we present means with standard deviations, as this makes comparison easier with those presented in POLYMOD [16] and in other social contact surveys [4]. As per previous studies, the sample was also weighted by 2/7 for weekends and 5/7 for weekdays to account for differences in sampling of weekend and weekend days and the difference between weekend and weekday contacts.

### Frequency and time spent with contacts

We explored types of behaviour with the frequency that participants met a contacted person, and with how long they spent with them. For this, we calculated the proportion of contacts that were physical, where a 2 metre distance was maintained, where a face-mask was used, and where they met outside. These were presented visually using stacked percentage bar charts. This approach was chosen as it allows for more direct comparison with the original POLYMOD paper [16] which explored duration and frequency with physical contact. We extend that analysis to include more pandemic specific behaviours.

### Contact matrices

For each country, we constructed age-stratified contact matrices for nine age groups (0 to 4, 5 to 11, 12 to 17, 18 to 29, 30 to 39, 40 to 49, 50 to 59, 60 to 69, and 70+ years old). For child participants and contacts, we did not record exact ages and therefore sampled from the reported age-group with a weighting consistent with the age distribution of contacts for the participants’ own age group, according to the POLYMOD survey methods [16]. Observations were weighted by 2/7 for weekends and 5/7 for weekdays. We fitted a negative binomial model censored to 50 per matrix cell, due to dispersion of the reported number of contacts, to calculate mean contacts between each participant and contact age groups. The value for censoring was chosen to be consistent and to ease comparison with previously published contact matrix estimates [8, 19]. To find the population normalised reciprocal contact matrix, we first multiplied the columns of the matrix by the mean-normalised proportion of the relevant country population in each age-group [16, 20]. Then we took the cross-diagonal mean of each element of the contact matrix. Finally, we divided the resulting symmetrical matrix by the population mean-normalised proportion of the country’s population in each age-group.

### Comparison to pre-pandemic and pandemic contact levels

We estimated the potential relative change in basic reproduction number R_0_ of an infection (that spreads along the contacts, assuming everyone would be susceptible to that infection) due to change in contact levels compared to pre-pandemic levels by calculating the ratio of the dominant eigenvalues of the CoMix matrices to those from POLYMOD, using the same approach as previously published [5]. Switzerland did not participate in the POLYMOD study and we therefore used an average of the eight countries for which data was collected to provide the pre-pandemic dominant eigenvalue for Switzerland. We also considered as an alternative the projected synthetic contact matrix for Switzerland from Prem et al[21], as a sensitivity analysis. Uncertainty for the ratios were provided by calculating the dominant eigenvalues from 1,000 bootstrap samples for the CoMix matrices for each country and the dominant eigenvalue from 1,000 bootstrap samples for the POLYMOD matrices for each country.

We further compared POLYMOD to the earliest estimates of contact levels during the 1st lockdown in the UK and BE. This estimate was not repeated for Switzerland and the Netherlands as data from children in these countries was not collected until later (December 2020).

### Comparison of post-pandemic and pandemic behaviours

We compared several of the measurements made during this final round of CoMix to those previously published from the prior rounds of the survey in order to frame the current findings in relation to those during the pandemic. We provide an exploratory but non-comprehensive comparison in order to reduce the burden for the reader to compare across multiple publications.

## Results

### Participant characteristics

Overall, we recorded observations on 7,477 participants who reported 74,534 contacts between 16 November 2022 and 6 December 2022 in the UK, Belgium, Netherlands, and Switzerland (Table 1). Just under 20% (1,336) were proxy respondents (i.e. the survey was completed by parents on behalf of children), and 6,141 were adults. The UK has the highest number of participants at 2,991, almost double the number of the other countries.

**Table 1:** Participants characteristics in the CoMix survey for each country.

| Category | Value | All | UK | BE | NL | CH |
| --- | --- | --- | --- | --- | --- | --- |
| All |  | 7,477 | 2,991 | 1,500 | 1,491 | 1,495 |
| Adult | N (%) | 6,141 (82.1%) | 2,488 (83.2%) | 1,200 (80.0%) | 1,215 (81.5%) | 1,238 (82.8%) |
| Child |  | 1,336 (17.9%) | 503 (16.8%) | 300 (20.0%) | 276 (18.5%) | 257 (17.2%) |
| Age group (Children) | 0-4 | 176 (15.4%) | 49 (14.0%) | 33 (11.3%) | 42 (16.3%) | 52 (21.4%) |
|  | 5-11 | 424 (37.1%) | 127 (36.4%) | 110 (37.7%) | 81 (31.4%) | 106 (43.6%) |
|  | 12-17 | 542 (47.5%) | 173 (49.6%) | 149 (51.0%) | 135 (52.3%) | 85 (35.0%) |
|  | Unknown | 194 | 154 | 8 | 18 | 14 |
| Age group (Adult) | 18-29 | 992 (17.0%) | 373 (16.6%) | 212 (17.7%) | 205 (17.3%) | 202 (16.9%) |
|  | 30-39 | 999 (17.1%) | 411 (18.3%) | 196 (16.3%) | 188 (15.8%) | 204 (17.1%) |
|  | 40-49 | 906 (15.5%) | 325 (14.5%) | 189 (15.8%) | 193 (16.3%) | 199 (16.7%) |
|  | 50-59 | 988 (16.9%) | 403 (17.9%) | 206 (17.2%) | 190 (16.0%) | 189 (15.8%) |
|  | 60-69 | 1,203 (20.6%) | 474 (21.1%) | 262 (21.8%) | 264 (22.2%) | 203 (17.0%) |
|  | 70+ | 743 (12.7%) | 263 (11.7%) | 135 (11.2%) | 147 (12.4%) | 198 (16.6%) |
|  | Unknown | 310 | 239 |  | 28 | 43 |
| Gender | Female | 3,781 (50.8%) | 1,564 (52.5%) | 733 (49.0%) | 759 (51.1%) | 725 (48.7%) |
|  | Male | 3,667 (49.2%) | 1,414 (47.5%) | 762 (51.0%) | 726 (48.9%) | 765 (51.3%) |
|  | Other | 29 | 13 | 5 | 6 | 5 |
| Household size | 1 | 1,508 (20.2%) | 538 (18.0%) | 295 (19.7%) | 339 (22.7%) | 336 (22.5%) |
|  | 2 | 2,520 (33.7%) | 1,062 (35.5%) | 473 (31.5%) | 487 (32.7%) | 498 (33.3%) |
|  | 3-5 | 3,292 (44.0%) | 1,323 (44.2%) | 699 (46.6%) | 638 (42.8%) | 632 (42.3%) |
|  | 6+ | 157 (2.1%) | 68 (2.3%) | 33 (2.2%) | 27 (1.8%) | 29 (1.9%) |
| Day of week | Sun | 1,796 (24.0%) | 533 (17.8%) | 317 (21.1%) | 656 (44.0%) | 290 (19.4%) |
|  | Mon | 856 (11.4%) | 357 (11.9%) | 41 (2.7%) | 111 (7.4%) | 347 (23.2%) |
|  | Tue | 1,663 (22.2%) | 676 (22.6%) | 570 (38.0%) | 26 (1.7%) | 391 (26.2%) |
|  | Wed | 1,704 (22.8%) | 950 (31.8%) | 256 (17.1%) | 322 (21.6%) | 176 (11.8%) |
|  | Thu | 848 (11.3%) | 419 (14.0%) | 117 (7.8%) | 234 (15.7%) | 78 (5.2%) |
|  | Fr | 366 (4.9%) | 24 (0.8%) | 132 (8.8%) | 88 (5.9%) | 122 (8.2%) |
|  | Sat | 244 (3.3%) | 32 (1.1%) | 67 (4.5%) | 54 (3.6%) | 91 (6.1%) |
| Risk perception (Adults) | Catching coronavirus | 441 (7.6%) | 139 (6.2%) | 89 (7.4%) | 122 (10.3%) | 91 (7.6%) |
| Strongly agree only | Serious illness from coronavirus | 554 (9.5%) | 187 (8.3%) | 133 (11.1%) | 159 (13.4%) | 75 (6.3%) |
|  | Spreading coronavirus to vulnerable people | 723 (12.4%) | 344 (15.3%) | 92 (7.7%) | 157 (13.2%) | 130 (10.9%) |
| Risk mitigation (Adults) | Face mask | 886 (14.4%) | 393 (15.8%) | 191 (15.9%) | 82 (6.7%) | 220 (17.8%) |
|  | Vaccinated | 5,269 (85.8%) | 2,196 (88.3%) | 1,044 (87.0%) | 1,044 (85.9%) | 985 (79.6%) |
|  | High risk | 1,468 (24.2%) | 423 (17.2%) | 347 (29.3%) | 372 (31.2%) | 326 (26.8%) |
| Symptoms (Adults) | Fever | 247 (4.2%) | 84 (3.7%) | 47 (3.9%) | 46 (3.9%) | 70 (5.9%) |
|  | Cough | 835 (14.3%) | 326 (14.5%) | 153 (12.8%) | 157 (13.2%) | 199 (16.7%) |
|  | Shortness of breath | 311 (5.3%) | 146 (6.5%) | 50 (4.2%) | 67 (5.6%) | 48 (4.0%) |
|  | Congestion | 892 (15.3%) | 315 (14.0%) | 180 (15.0%) | 198 (16.7%) | 199 (16.7%) |
|  | Sore throat | 554 (9.5%) | 199 (8.8%) | 118 (9.8%) | 106 (8.9%) | 131 (11.0%) |
|  | Fatigue or tiredness | 551 (9.4%) | 237 (10.5%) | 97 (8.1%) | 115 (9.7%) | 102 (8.5%) |
|  | Any symptoms | 2,324 (39.9%) | 872 (38.8%) | 462 (38.5%) | 473 (39.8%) | 517 (43.3%) |
| Employed (Adults) | Full time | 1,772 (68.2%) | 677 (69.1%) | 379 (77.3%) | 331 (59.4%) | 385 (67.3%) |
|  | Part time | 682 (26.2%) | 251 (25.6%) | 87 (17.8%) | 192 (34.5%) | 152 (26.6%) |
|  | Self employed | 145 (5.6%) | 52 (5.3%) | 24 (4.9%) | 34 (6.1%) | 35 (6.1%) |
| Work open (Adults) | Closed | 297 (9.1%) | 148 (11.2%) | 48 (8.0%) | 51 (7.5%) | 50 (7.5%) |
|  | Open | 2,980 (90.9%) | 1,179 (88.8%) | 552 (92.0%) | 630 (92.5%) | 619 (92.5%) |
| Attended work (Adults) | Yes | 1,632 (61.6%) | 598 (60.0%) | 307 (61.5%) | 299 (52.3%) | 428 (73.8%) |

The age distributions were broadly similar across the four countries, with Switzerland the most different with slightly more over 70s and fewer 60-69, and more 5-11s and fewer 12-17 year olds. There were 3,781 (50.8%) females and 3,667 (49.2%) males, with a similar roughly equal split in all countries. The majority of households consisted of 3-5 people in total with less than 2.5% of participants in any country being in a household size of six or more. Contact data was collected on every day of the week for all countries, though some days had lower participation such as 24 (0.8%) and 32 (1.1%) responses in the UK on Friday and Saturday, and 41 (2.7%) in Belgium on Monday, and 26 (1.7%) in the Netherlands on Tuesdays.

### Risk Perception

Overall, 7.6% of the sample (ranging from 6.2% in the UK to 10.3% in the Netherlands) strongly agreed that they were at risk of catching coronavirus and 9.5% strongly agreed that they were at high risk of severe disease if they did catch coronavirus (ranging from 6.3% in Switzerland to 13.4% in the Netherlands). A slightly higher fraction (12.4%) strongly agreed that they were likely to spread the virus to someone vulnerable, varying from 7.7% in Belgium to 15.3% in the UK.

### Risk Mitigation

Only 14.4% of participants reported wearing a facemask on the previous day. The Netherlands had the lowest with 6.7% participants wearing a facemask and Switzerland the highest with 17.8% (Table 1). Self-reported vaccination in adults was similar for each country at around 85% vaccinated. The UK had the lowest percentage of people self-reporting as being high risk at 17.2% versus 31.2% in the Netherlands.

### Symptoms

Nearly 40% of participants reported at least one of the following symptoms: fever or chills, cough, shortness of breath (or difficulty breathing), fatigue (or extreme tiredness), muscle or body aches or headache, congestion (or runny nose), and sore throat.

### Employment

About 43% of adult participants were employed, though this includes individuals who may be retired as unemployed in the denominator. Of those that were employed, the majority (between 60 to 80%) in each country were in full time employment, and around 5% were self employed. For those in employment the vast majority (∼90%) reported their workplaces were open and around two thirds attended work in person on the day they made their contacts (Table 1).

### Mean Contacts by country and setting

Participants from the Netherlands recorded considerably more contacts than the other three countries with 9.9 (95% CI 9.0 to 10.8) contacts per person per day, as compared to 6.5 (95% CI 6.0 to 7.0) contacts in UK, 6.7 (95% CI 6.0 to 7.3) in Belgium and 6.0 (95% CI 5.4 to 6.6) in Switzerland. (Table 2). This pattern was also seen for adults and children (8.8, 95% CI 7.9 to 9.8 for adults; 14.8, 95% CI 12.6 to 16.8 for children in the Netherlands). As well as overall contacts, we measured contacts for the four settings of home, work, school, and other. Contacts at home were very similar between the countries, with an average of about 1.5 contacts per person per day recorded, which is consistent with the household sizes seen in Table 1 (a mean of 2.6 overall for the study). Contacts at work for adults were lowest in the UK (a mean of 1.5 contacts recorded per person per day, 95% CI 1.2 to 1.9) and highest in the Netherlands at 3.3 contacts per person per day (95% CI 2.7 to 4.0). Other contacts (mostly in social settings) were also lowest in the UK at 1.6 per person per day (95% CI 1.4 to 1.9) and highest in the Netherlands at 3.3 recorded per person per day (95% CI 2.7 to 4.0).

**Table 2:** Mean contacts by country and setting.

| Category | Setting | UK | BE | NL | CH |
| --- | --- | --- | --- | --- | --- |
| All participants |  | Mean (95% CI*) |  |  |  |
|  | All | 6.5 (6.0 to 7.0) | 6.7 (6.0 to 7.3) | 9.9 (9.0 to 10.8) | 6.0 (5.4 to 6.6) |
|  | Home | 1.5 (1.5 to 1.6) | 1.6 (1.5 to 1.6) | 1.6 (1.5 to 1.7) | 1.5 (1.4 to 1.5) |
|  | Work | 1.4 (1.1 to 1.7) | 1.7 (1.3 to 2.1) | 2.8 (2.3 to 3.3) | 1.6 (1.3 to 1.9) |
|  | School | 2.2 (1.9 to 2.6) | 1.7 (1.3 to 2.1) | 3.0 (2.5 to 3.5) | 1.1 (0.8 to 1.4) |
|  | Other | 1.6 (1.4 to 1.9) | 2.2 (1.9 to 2.6) | 3.4 (3.0 to 4.0) | 2.2 (1.9 to 2.7) |
| Adults |  |  |  |  |  |
|  | All | 5.4 (5.0 to 5.9) | 5.5 (4.8 to 6.2) | 8.8 (7.9 to 9.8) | 5.3 (4.8 to 5.9) |
|  | Home | 1.4 (1.3 to 1.4) | 1.3 (1.3 to 1.4) | 1.4 (1.3 to 1.5) | 1.3 (1.2 to 1.4) |
|  | Work | 1.5 (1.2 to 1.9) | 2.1 (1.7 to 2.7) | 3.3 (2.7 to 4.0) | 1.9 (1.5 to 2.3) |
|  | School | 1.2 (1.0 to 1.5) | 0.5 (0.3 to 0.8) | 1.9 (1.4 to 2.4) | 0.5 (0.3 to 0.7) |
|  | Other | 1.6 (1.4 to 1.9) | 2.0 (1.6 to 2.3) | 3.3 (2.7 to 4.0) | 2.1 (1.7 to 2.5) |
| Children |  |  |  |  |  |
|  | All | 11.1 (9.4 to 12.7) | 10.4 (8.7 to 12.3) | 14.8 (12.6 to 16.8) | 9.1 (7.4 to 11.1) |
|  | Home | 2.2 (2.1 to 2.3) | 2.2 (2.1 to 2.4) | 2.6 (2.4 to 2.8) | 2.1 (1.9 to 2.3) |
|  | Work | 0.9 (0.4 to 1.5) | 0.1 (0.0 to 0.3) | 0.3 (0.1 to 0.4) | 0.4 (0.2 to 0.7) |
|  | School | 6.7 (5.3 to 8.1) | 5.5 (4.3 to 6.9) | 8.1 (6.6 to 9.6) | 4.0 (2.8 to 5.2) |
|  | Other | 1.7 (1.2 to 2.3) | 3.1 (2.2 to 4.0) | 4.0 (3.1 to 4.9) | 3.1 (2.1 to 4.2) |

\*Bootstrapped mean and 95% percentage confidence interval from 1,000 samples. Sample weighted by 2/7 for weekends and 5/7 for weekdays.

### Frequency and time spent with contacts

Higher frequency contacts (1-2 days) were more likely to include physical touch (> 50%) compared to less frequent contacts (e.g. never met <25%) (Figure 1). Similarly, physical contact was more likely for those spending 4 hours or more with a contact, with the proportion of physical contacts observed in the data reducing as the duration of contact reduced (Figure 2).

**Figure 1:**
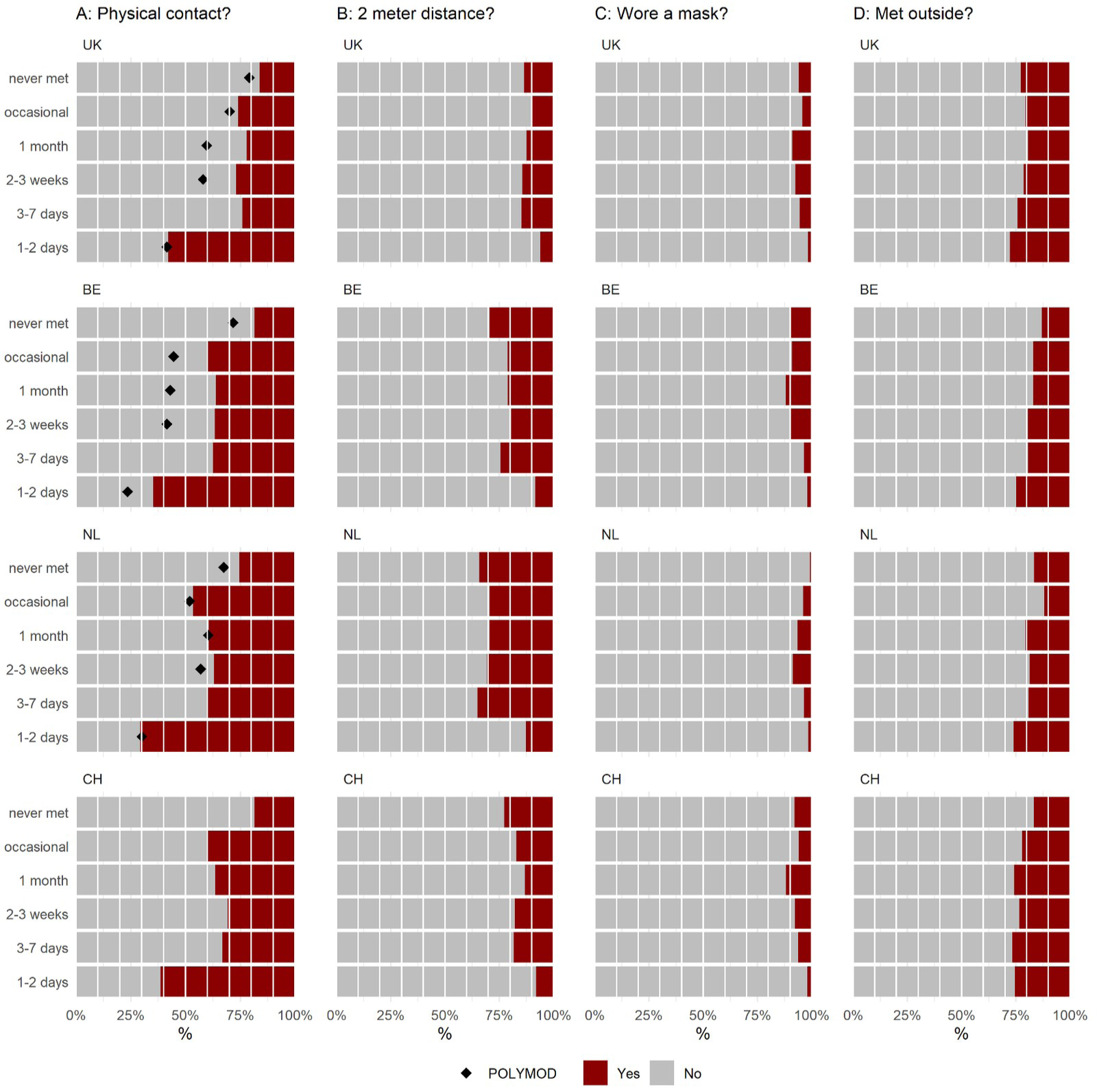
Frequency of meeting the contacts for each country by: **A**. Whether it was a physical contact or not, **B** whether they were further than 2 metres from the contact, **C** Whether they wore a mask when meeting the contact, **D** whether they met outside when meeting the contact

**Figure 2:**
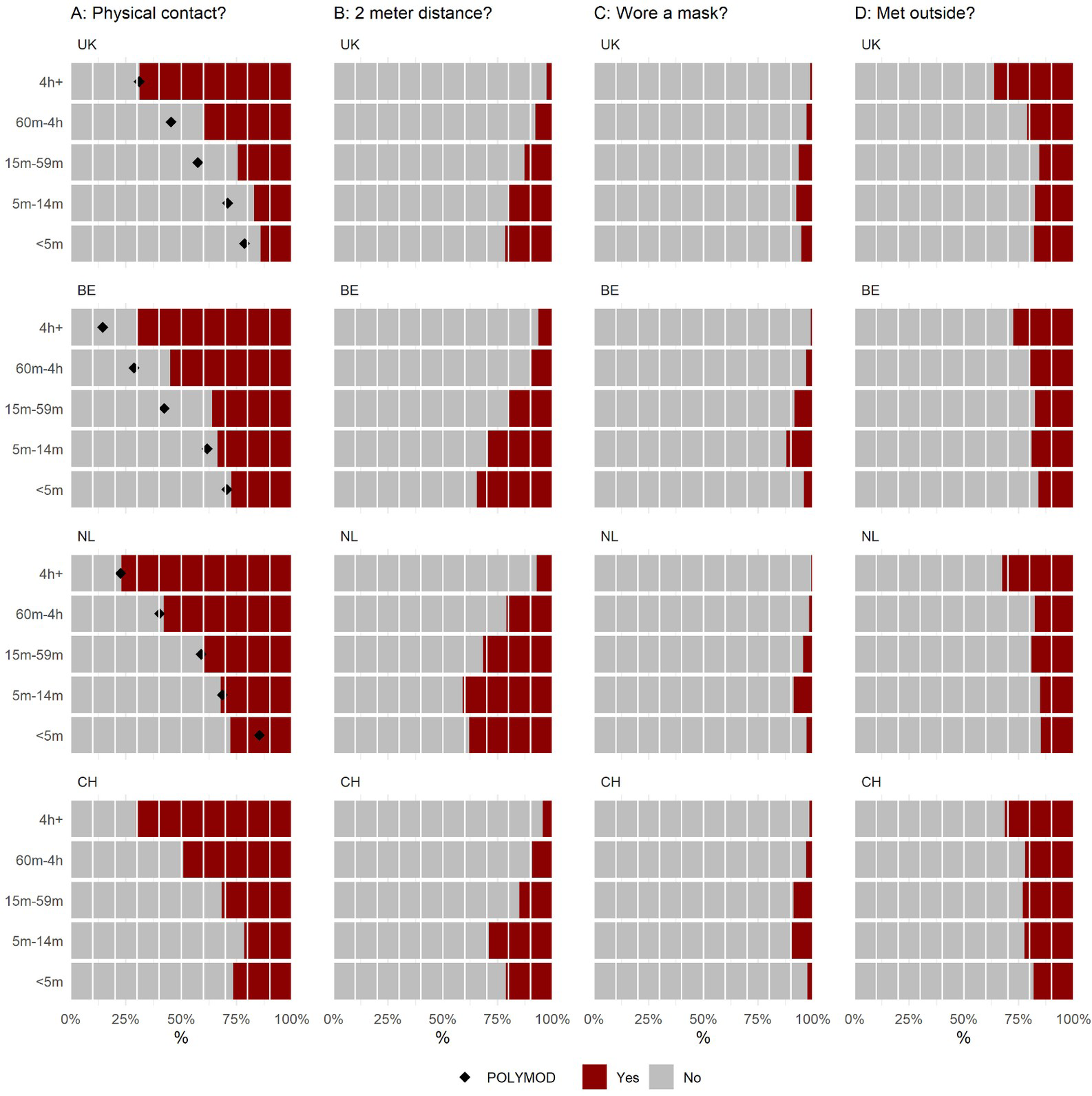
Time spent with contacts for each country by: **A**. Whether it was a physical contact or not, **B** whether they were further than 2 metres from the contact, **C** Whether they wore a mask when meeting the contact, **D** whether they met outside when meeting the contact

The percentage of contacts met every 1-2 days that were physical was similar to those seen in POLYMOD for the UK, Netherlands, and Belgium (Figure 1). Though for those meeting less often, the percentage of physical contacts appears lower than POLYMOD for the UK and Belgium but similar for the Netherlands (Figure 1). The patterns were somewhat consistent for time spent with contacts and percentage of physical contact, the Netherlands had near identical percentages, Belgium had slightly lower for all but the shortest durations of contacts, and the UK had slightly lower for all but the longest duration of contacts (Figure 2).

The percentage of participants staying at least 2 metres away was slightly higher in the Netherlands though still less than 25% for all countries, with only those who were met every 1-2 days being less likely to wear a mask compared to when meeting a less frequent contact (Figure 1). Maintenance of a two metre distance appears to be more common for shorter interactions (Figure 2),

Mask wearing was infrequent (<15%) in all countries and for all types of contact, with participants less likely to wear a mask when meeting someone often and for shorter periods (<5m) or longer (4h+) periods of time (Figures 1 and 2).

The fraction of contacts who met outside were similar for all frequency of contact and across the four countries (Figure 1). There was a slight trend (in each country) for longer-duration contacts to have occurred outside (Figure 2).

### Mean contacts by characteristics

#### Age, Gender, Households size

The reported mean contacts for school-aged (5-11 and 12-17 years of age) in the UK and Netherlands were similar at around 14 contacts per person per day, whereas Belgium and Switzerland were lower with both at around 10 contacts (Table 3). This pattern was different amongst adults, with the UK reporting the lowest levels of contacts in most adult age groups. Young adults (18-29 years old) in Belgium and the Netherlands reported the highest mean contact rates (7.6 and 10.4 per person per day, respectively, as compared to 4.8 in the UK and 5.9 in Switzerland).

**Table 3:**
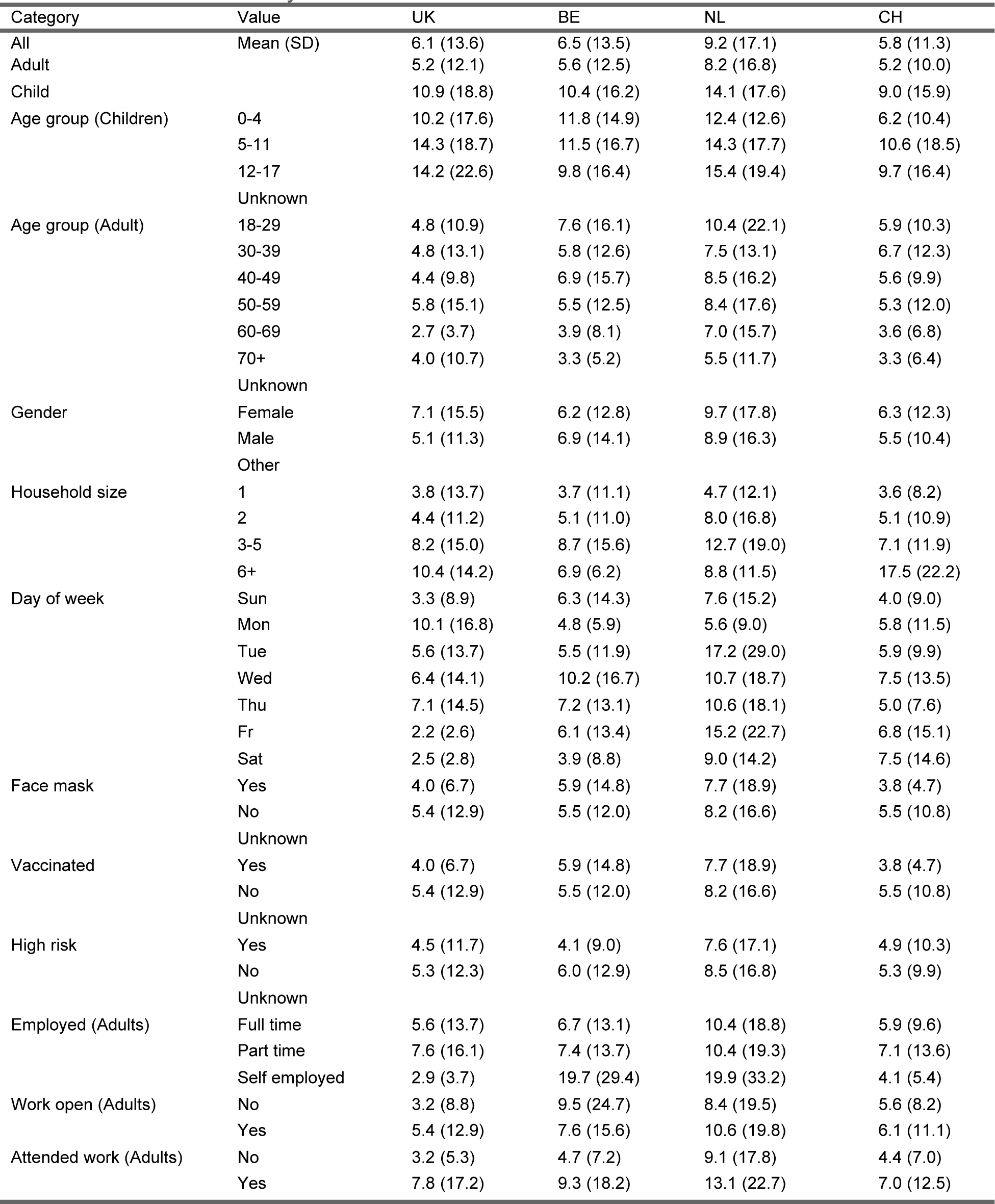
Mean contacts by characteristics.

Females generally reported more contacts than males, though this pattern was not consistent in each country. As expected, household size was positively correlated with the number of reported contacts with some slight departures from this pattern in Belgium and the Netherlands.

#### Day of the week

Contacts by day suggest a strong weekend effect for all countries, with far lower contacts on the weekend and also on a day either side of the weekend for the UK (Friday) and Belgium and the Netherlands (Monday) (Table 3).

#### Risk mitigation

Those who reported wearing a facemask tended to report fewer contacts in all countries other than Belgium. Those self reporting as high risk reported lower contacts across all four countries. Those who were vaccinated tended to report fewer contacts than those who said they had not been vaccinated (except for in Belgium), though it should be stressed that this is a univariate analysis and the unvaccinated tended to be younger in age.

#### Employment

Number of contacts were highest for employed people in the Netherlands, with self employed people in Belgium and the Netherlands reporting about 20 contacts per person per day. With the vast majority of workplaces being open now, contacts still tended to be higher for people whose workplace was open. As expected, there was still a considerable difference in the mean contacts for those who attended work versus those who did not.

### Contact matrices and changes in pre-pandemic and post-pandemic R_0_

Contact matrices were similar across the four nations, with high rates of recorded contacts along the leading diagonal (suggesting that contact is age-assortative) and the highest rates of recorded contacts being for children (Figure 3A). The Netherlands had the highest levels of contacts overall. There were comparatively high levels of contact between over 70s in all countries, except Belgium.

**Figure 3:**
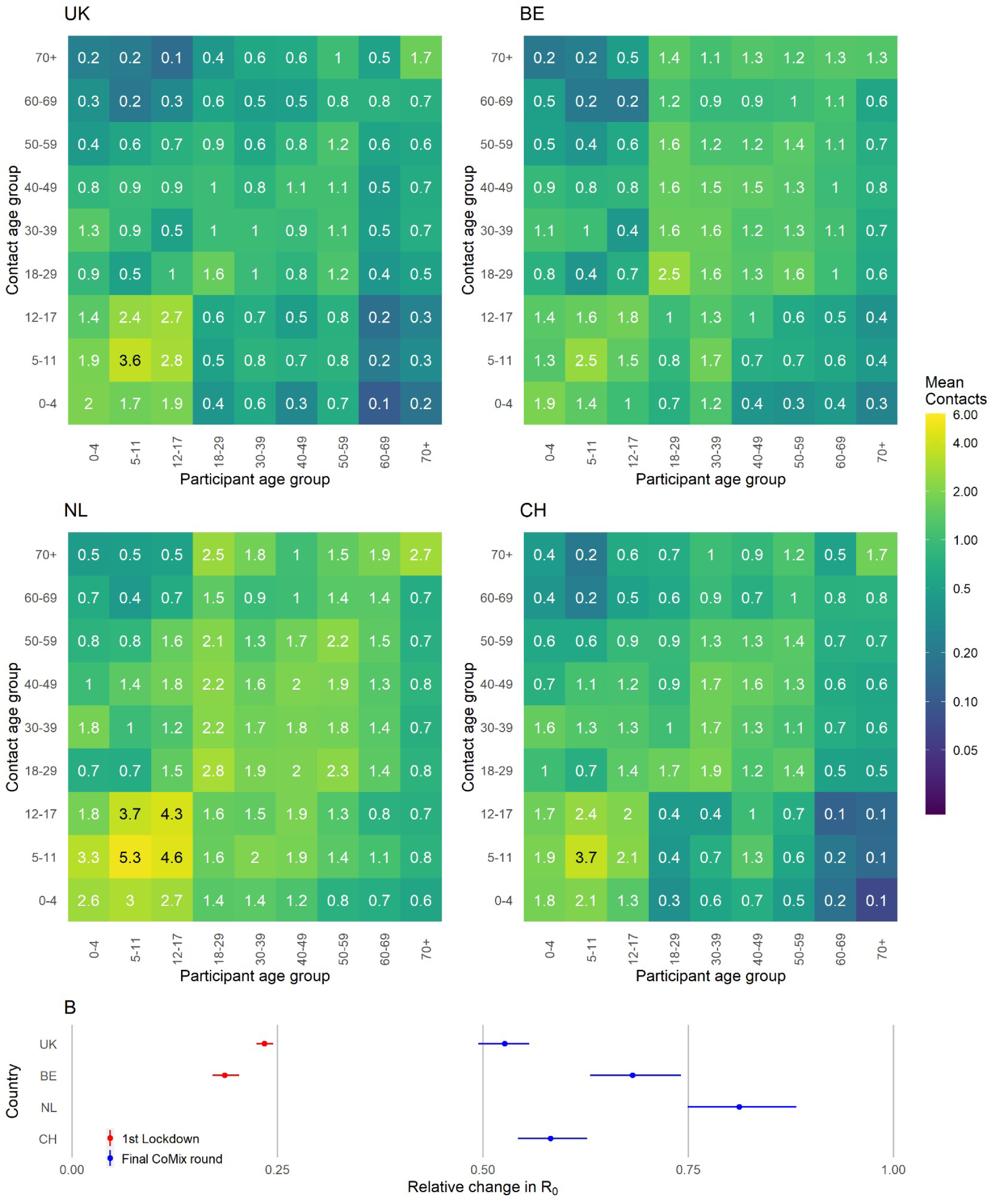
**A**: Contact matrices for each country. **B:** Points show relative change in R_0_ (compared to POLYMOD) based on the dominant eigenvalues of contact matrices.

Using the next-generation approach [22], these contact matrices can be used to estimate R_0_ for close-contact infections spread through physical or conversational contacts (as measured here), assuming that everyone is susceptible to infection. The relative change in R_0_ for reported contacts, compared to contacts at pre-pandemic levels (as measured in the POLYMOD study) is shown in Figure 3B (Table S1). The reduction in contacts, compared with POLYMOD, would lead to a significant reduction in the reproduction number R_0_ in each of the four countries, with the UK’s R_0_ being roughly half of pre-pandemic levels and the Netherlands about 80% of the pre-pandemic level (with the other two countries being intermediate). For context, Figure 3B also shows the relative reduction in R_0_ during the first lockdown in the UK, which was 25% of the pre-pandemic level and Belgium which was 20% of pre-pandemic levels (Figure 3B and Table S1) [6]. Using the projected contact matrix for Switzerland by Prem et al. [21] instead of the POLYMOD average leads to comparable results (Figure SI1).

### Comparison of post-pandemic and pandemic behaviours and contacts

In this section comparisons are made between the results from this round and those found for the UK, Netherlands, Belgium, and Switzerland in the analysis of the 2-year 21 country study by Wong et al [9].

#### Risk Perception, Mitigation, and Symptoms

It appears that risk perceptions may have slightly changed since the pandemic with participants reporting a slightly higher belief that they will catch coronavirus (6-10% versus 4-5% in Wong), and a lower concern that it would be a serious illness for them (6 to 13% versus 14 to 25% in Wong), or that they will pass it on to someone considered vulnerable (8 to 15% versus 19 to 26% in Wong).

The percentage of individuals wearing facemasks (7 to 18%) were considerably lower than levels measured in each individual country over the course of the pandemic (UK, 58.2%, BE 61.4%, NL 34.3%, CH 76.7%) [9].

The percentage of participants reporting at least one symptom (38 to 43%) was quite a bit higher than reported over the two years of the pandemic which was between 21 to 26% [17]. (Table 1 in this paper versus Table 1 in Wong et al [9]).

### Contacts

The mean contacts measured in this survey were somewhat higher at between 6-10 for the mean contacts for the four countries compared to 3-4 contacts per day measured during the pandemic [9]. Apart from a general increase in the level of contacts the main change appears to be in those 70+, an age group with very few contacts made during the pandemic, especially in the UK (see Figure 6A in Gimma et al [8]). In contact matrices measured during the pandemic in the UK, those aged 70 or older never had more than 1 contact on average with those also aged 70+ and less than 0.4 for contacts with other age groups. In contrast, we estimate a value of 1.7 for 70+ year olds mixing with 70+ year olds and values as high as 0.7 for mixing with other age groups.

## Discussion

We estimate that contact levels have increased compared to those measured during the pandemic but still remain lower than those measured prior to the pandemic. These reduced levels are likely to have a big impact on transmission with a reduction of R_0_ of between 20% to 50% compared to pre-pandemic levels across the four nations. The consequences of this change in behaviour extends well beyond Covid and would be expected to have an impact on a range of infections that are spread person-to-person.

The use of facemasks has dropped considerably compared to the levels measured during the pandemic. We estimated around 15% of people wore a face mask on the day of the study across the four countries which is considerably lower than the 64% average observed during the pandemic across 21 European countries [17].

Contacts amongst the individuals over the age of 70 were consistently low during the pandemic and we observed a bounce back in the number of contacts over 70s make especially in the social setting.

Contact patterns were broadly similar across the four countries, with the Netherlands generally reporting a higher level of contacts. The patterns of the frequency of contacts, whether they’re physical or not, and the duration of contacts were somewhat similar to those seen prior to the pandemic.

We also observed that the proportion of individuals who think they are likely to get Covid was higher than those measured during the pandemic but there has potentially been a shift that it is considered less serious for them and there is less concern about giving it to someone vulnerable. Given the relationship between perceived severity and contacts measured during the pandemic, this is one of the potential explanation for the increase in contacts that we observed [23].

The CoMix study was nearly identical in the four countries, with the same questionnaire (apart from translation issues) and a similar sampling frame, and collected by the same survey organisation at the same calendar time. The study design was also the same as those used for the previous rounds of CoMix which allows for more straightforward comparison to the estimate calculated during the pandemic. We also structured our analyses to be consistent with previous analyses conducted for POLYMOD and CoMix.

A difficulty of our study design is that it is retrospective (individuals were asked about their contacts on the previous day), so may miss contacts, particularly those that would be short lasting. Furthermore, the children’s contacts are a proxy with parents reporting on behalf of those under-18. We also allow individuals to estimate mass contacts that they were unable to report individually, which results in skewed distributions of contacts and is why a maximum threshold value of 100 contacts per person is used for estimates of the mean.

This research provides a snapshot picture of contacts in four European nations during the return to post-pandemic patterns of behaviour. We have measurements that are higher than those seen during the pandemic but are still considerably lower than those prior to the pandemic. It may be that the huge changes we saw during the pandemic are not over, and it will be important to monitor changes in contacts that may occur over the coming years.

It appears that the pandemic, at least in terms of behaviour, is ending very slowly and we are seeing a long return to contact level prior to 2019. However, it could be that we may never return to the levels of contacts seen before the pandemic. The changes in work patterns, and behaviour may have resulted in long-lasting impacts with implications on the epidemiology of a wide range of infections, as well as on important societal and economic outcomes.

## Conclusions

Despite the number of contacts being higher compared to pandemic levels, we are not back to the levels seen prior to the pandemic. The Netherlands and Belgium appear closer to pre- pandemic levels with the UK further behind. These divergences between countries may represent long-term changes and measuring the level of social interactions in the years to come will allow this to be assessed. Pandemics may not end with a bang but perhaps rather a slow and cautious trudge back to newly considered risky behaviour that was previously part of everyday life.

## Data Availability

The code and data used to conduct these analyses are found at https://github.com/jarvisc1/cmix_post_pandemic.

## Abbreviations

CI: confidence interval
UK: United Kingdom
CH: Switzerland
BE: Belgium
NL: Netherlands

## Declarations

### Authors’ contributions

WJE, and CIJ designed the CoMix contact survey. CIJ conceived of and planned the analysis. CIJ, PC, JAB, JDM, PB, NH, CLA, JW, CF, and WJE provided comments and discussions on analytical methods. CIJ, JDM, and PC conducted the analysis. CIJ wrote the first draft of the manuscript with feedback from all other authors.

## Acknowledgements

We acknowledge support from the European Centre for Disease Prevention and Control (ECDC) in setting up the collaborations between the Epipose consortium, and universities and public health institutions in all other countries. We gratefully acknowledge the tremendous efforts put in all the steps by the EpiPose consortium, its collaborators and Ipsos.

We would also like to thank the team at Ipsos who have been excellent in running the survey, collecting the data, and allowing for this study to, happen at a rapid speed. We acknowledge the exceptional project management support given by Sarah Vercruysse, Bieke Vanhoutte and Anna Carnegie.

## Consent for publication

Not applicable. We do not report individual patient data.

## Additional Files

**Table S1:** Estimates of R0 from Figure 3B.

| country | period | $R_0$ | 2.5% | 97.5% |
| --- | --- | --- | --- | --- |
| UK | Final CoMix round | 0.526 | 0.494 | 0.557 |
| BE | Final CoMix round | 0.682 | 0.631 | 0.741 |
| NL | Final CoMix round | 0.813 | 0.749 | 0.881 |
| CH | Final CoMix round | 0.583 | 0.543 | 0.627 |
| UK | 1st Lockdown | 0.234 | 0.225 | 0.244 |
| BE | 1st Lockdown | 0.186 | 0.171 | 0.203 |

**Figure S1:**
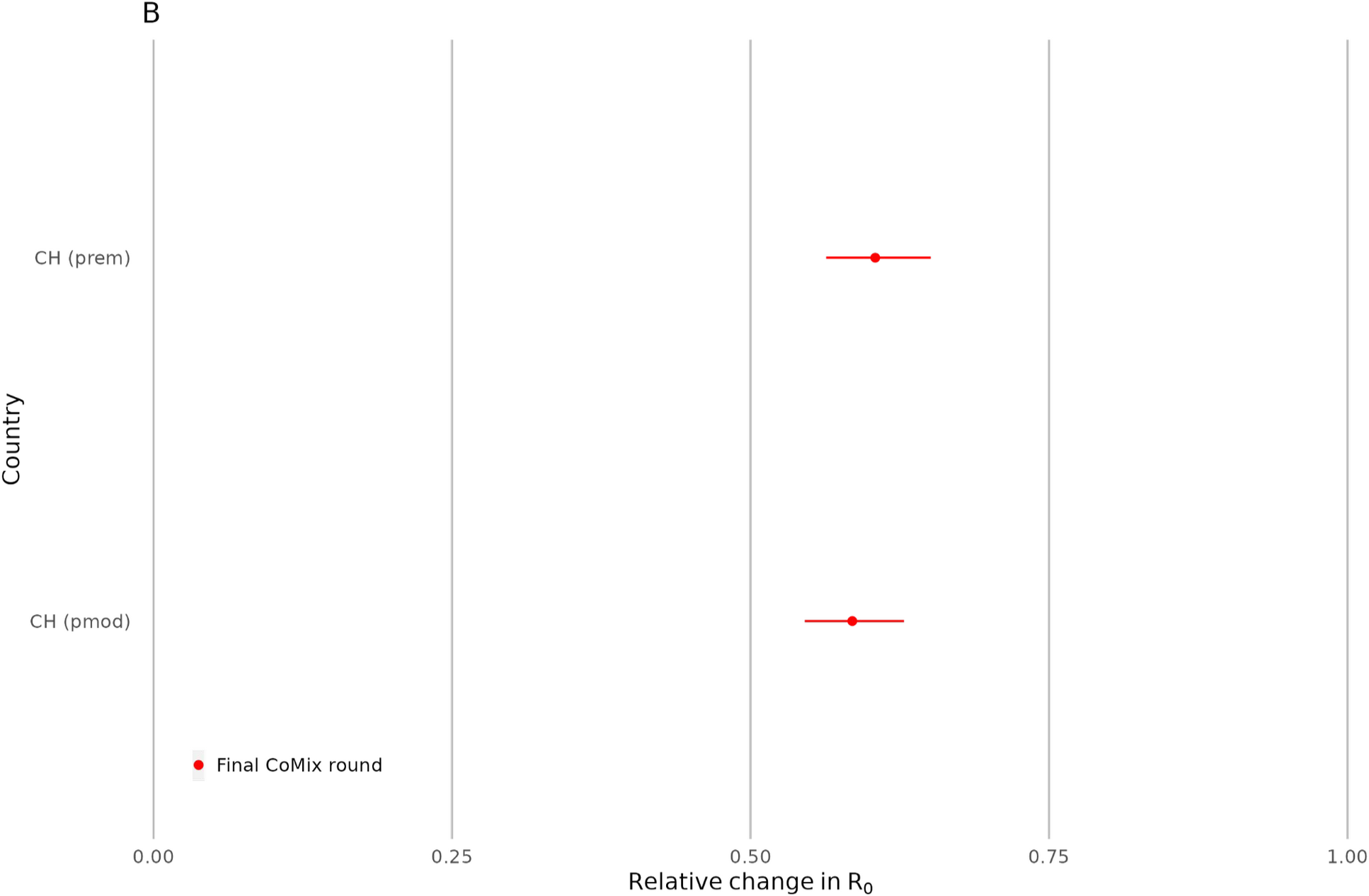
Reduction in R_0_ for Switzerland when using different baseline: Average of the Polymod data (“CH (pmod)” bottom row) vs projected contact matrix from Prem et al [21] (“CH (prem)”, bottom row).

